# Genetic variants in the *SHISA6* gene are associated with delayed cognitive impairment in two family datasets

**DOI:** 10.1101/2021.07.02.21259940

**Authors:** Jairo Ramos, Laura J. Caywood, Michael B. Prough, Jason E. Clouse, Sharlene D. Herington, Susan H. Slifer, M. Denise Fuzzell, Sarada L. Fuzzell, Sherri D. Hochstetler, Kristy L. Miskimen, Leighanne R. Main, Michael D. Osterman, Owen Laframboise, Andrew F. Zaman, Patrice L. Whitehead, Larry D. Adams, Renee A. Laux, Yeunjoo E. Song, Tatiana M. Foroud, Richard P. Mayeux, Peter St. George-Hyslop, Paula K. Ogrocki, Alan J. Lerner, Jeffery M. Vance, Michael L. Cuccaro, Jonathan L. Haines, Margaret A. Pericak-Vance, William K. Scott

## Abstract

**Background:** Studies of cognitive impairment (CI) in Amish communities have identified sibships containing multiple CI and cognitively unimpaired (CU; unaffected after age 75) individuals. We hypothesize that these CU individuals may carry protective alleles delaying age at onset (AAO) of CI, preserving cognition in older age despite increased genetic risk. As well, the genetic and cultural isolation in the Amish since the early 1800s may have reduced the complexity of the genetic architecture of CI, increasing the power to detect protective alleles in this population. With this in mind we conducted a genome-wide study (GWAS) to identify loci associated with AAO of CI in a sample of Amish adults over age 75.

**Methods:** 1,522 individuals aged 43-99 (mean age 73.1, 42% men) screened at least once for CI using the Modified Mini-Mental State exam (3MS) were genotyped using Illumina chipsets. Genotypes were imputed for 7,815,951 single nucleotide variants (SNV) with minor allele frequency (MAF) > 1%. The outcome studied was age, defined as 1) age at the first 3MS result indicating impairment (AAO; 3MS <87; 362 CI individuals) or 2) age at last normal exam (3MS >=87, 1,160 CU individuals). Cox mixed-effects models examined association between age and each SNV, adjusting for sex and familial relationships. To replicate genome-wide significant findings, SNVs in a 1 Megabase region centered on the peak SNV were examined for association with age using these same methods in the NIA-LOAD family study dataset (1,785 AD cases, 1,565 unaffected controls, mean age 73.5.

**Results:** Three SNV were significantly associated (p<5 x 10-8) with AAO in the Amish, on chromosomes 6 (rs14538074; HR=3.35), 9 (rs534551495; HR=2.82), and 17 (rs146729640; Hazard Ratio (HR)=6.38). Each region found the common allele associated with later AAO. Replication analysis detected association at rs146729640, with nominal statistical significance (HR=1.49, p=0.02).

**Conclusions:** The replicated genome-wide significant association with AAO on chromosome 17 suggest this may be novel locus associated with delayed onset of AD. The associated SNP is located in the *SHISA6* gene, which is involved in post-synaptic transmission in the hippocampus and is a biologically plausible candidate gene for AD.

## INTRODUCTION

Dementia, which includes Alzheimer disease (AD), cerebrovascular, Lewy body, and mixed dementias (Hale et al.,2020), has a tremendous effect on the entire family, since the largest proportion, at least 60 % in the U.S., of caregivers are spouses, children and children-in-law (Brodaty & Donkin, 2009). Dementia also carries a heavy emotional and financial toll, as people can live for many years cognitively impaired, requiring high levels of care with annual estimated costs in the U.S. in 2010 between $159 billion and $215 billion (Hale et al.,2020) (Hurd et al., 2013). Understanding the factors that influence the age at which cognitive impairment begins is foundational in developing approaches toward reducing the prevalence and progression of disease. Much effort has been made to identify genetic factors that increase the risk and result in earlier onset of dementia, and dozens of risk variants have been identified. The search for protective variants, including those that delay age at onset, has been less common. Studies of time to onset of dementia symptoms, employing survival analysis approaches, offer a potentially powerful means of identifying protective variants in age related neurodegenerative diseases that shorten the duration of cognitive impairment (CI) by delaying onset of symptoms.

The complex genetic architecture of dementia and AD has complicated the search for both risk and protective variants. Reproducibility of genetic association is influenced by locus heterogeneity, smaller sample sizes, and population differences. Some of these limitations are addressed by large-scale consortium approaches to genome-wide association studies (GWAS; Kunkle et al., 2019). A second strategy to improve power and reduce heterogeneity is to study a genetically homogenous population such as the Amish (Hahs et al., 2006). The Amish in the United States live in relatively genetically isolated communities (Hahs et al., 2006); members of these communities have participated in longitudinal studies of aging, cognition, and cognitive impairment for over two decades (Pericak-Vance et al., 1996) (Hahs et al., 2006) (Edwards et al., 2011) (Cummings et al., 2012) (Edwards et al., 2013) (D’Aoust et al., 2015) (Ramos et al., 2021) which have identified sibships with both individuals with CI and siblings that are cognitively unimpaired (CU). Our hypothesis is that individuals that are CU carry genetic variants that delay AAO of CI in spite of having an increased risk due to advanced age and family history of CI in a sibling.

In this study we performed a GWAS of AAO of CI in 1,522 people from the Amish communities living in Ohio and Indiana. Genome-wide significant results were examined for replication in an independent sample of 3,350 individuals from the NIA-LOAD family study of AD. A replicated association on chromosome 17 was identified, supporting the hypothesis that genetic variants delay the development of cognitive impairment in individuals who are CU despite elevated risk due to age and family history of dementia.

## METHODS

### Study Populations and Clinical Assessment

A description of the sample of 1,522 Amish used in this study is provided in Table 1. All participants were of European descent, with an average age of 73 at the last examination (range: 43 to 99). Women were 58.3% of the sample. Individuals were residents of Amish communities in Ohio (Holmes County) and Indiana (Adams, Elkhart, and LaGrange counties) and recruited as previously described by Pericak-Vance et al. (1996), Hahs et al. (2006), Edwards et al. (2013), Edwards et al. (2014) and Ramos et al. (2021). This is part of a population-based study of aging and age-related phenotypes such as AD performed by researchers from the University of Miami and Case Western Reserve University after Institutional Review Board (IRB) review and approval.

**Table 1.**
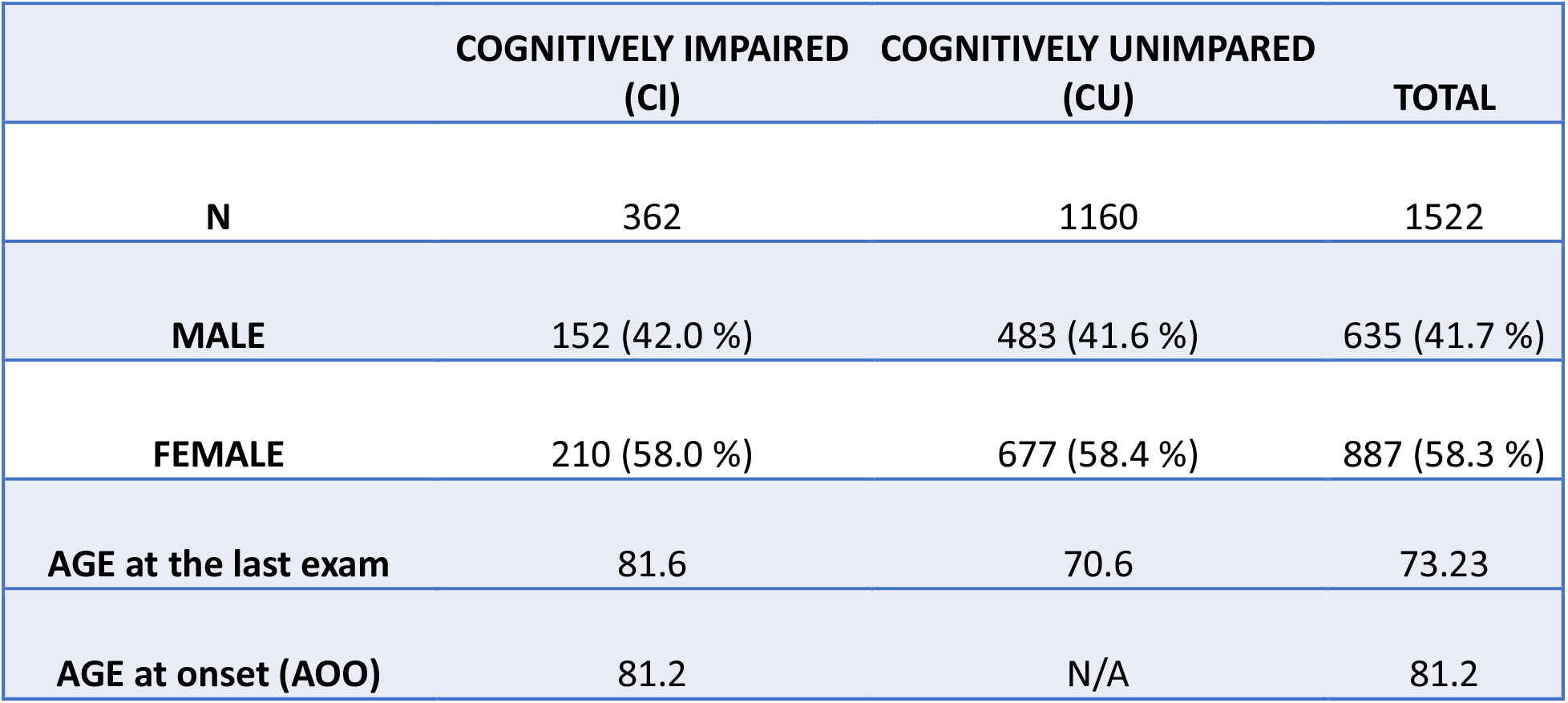
Demographic characteristics of the 1,522 Amish individuals included in the study. About 24% of the sample was cognitively impaired at the last examination. Cognitively impaired individuals were older at the last examination, and the two groups had similar proportions of men and women.

All Amish participants were evaluated for cognitive function at least once using the Modified Mini-Mental State exam (3MS, Teng & Chui, 1987). DNA was extracted from a blood sample obtained at this baseline examination, which also included an assessment of family history of dementia or AD, a medical history of common chronic diseases, assessments of self-reported physical function and objective physical function, and basic sociodemographic information. The average number of clinical examinations was 1.4 (range: 1 to 5).

Individuals with education-adjusted 3MS scores below 87 at baseline or at a follow-up visit were classified as cognitively impaired (CI). Those scoring 87 and above at all visits were considered cognitively unimpaired (CU). Age at onset of cognitive impairment (AAO of CI) was determined as the age at the examination when the participant was observed impaired for the first time. For individuals with CI at baseline, this may overestimate AAO of CI. For survival analysis, the outcome was defined as AAO of CI for those with cognitive impairment, and age at last examination for those classified as CU. The outcome of interest here was *CI-free time*, rather than the development of dementia, and therefore we chose a broad definition of CI (including AD and other types of dementia) as the event, rather than narrower definitions of AD. This allows the identification of loci protective against a wider range of dementia phenotypes, at the possible expense of greater heterogeneity of effects and lower power if the loci are specific to AD. We therefore attempted to replicate our findings in a second family dataset with individuals meeting this more specific AD case definition and their unaffected relatives.

For external replication we used the family-based National Institute on Aging Genetics Initiative for Late-Onset Alzheimer’s Disease (NIA-LOAD) (Wijsman et al., 2011) (Tosto et al., 2017) dataset composed of 3,350 subjects (1,785 late onset AD (LOAD) cases and 1,565 controls neurologically evaluated as unaffected). Subjects were evaluated using standard research criteria for the diagnosis of LOAD as established in McKhann et al. (1984) including the Clinical Dementia Rating score (Hughes et al., 1982). Cases were defined as those individuals with definite or probable LOAD diagnosis as per guidelines in McKhann et al. 1984, while the age at onset (AAO) was established as the age at which the family first observed memory problems or the age at first exam where cognitive impairment was noted. For controls the age used was the age at last examination when the absence of dementia was determined (McKhann et al. 1984) (Lee et al., 2008). The demographics of the sample are provided in Table 2. More details about the sample are described in Lee et al. (2008) and Kunkle et al. (2019).

**Table 2.**
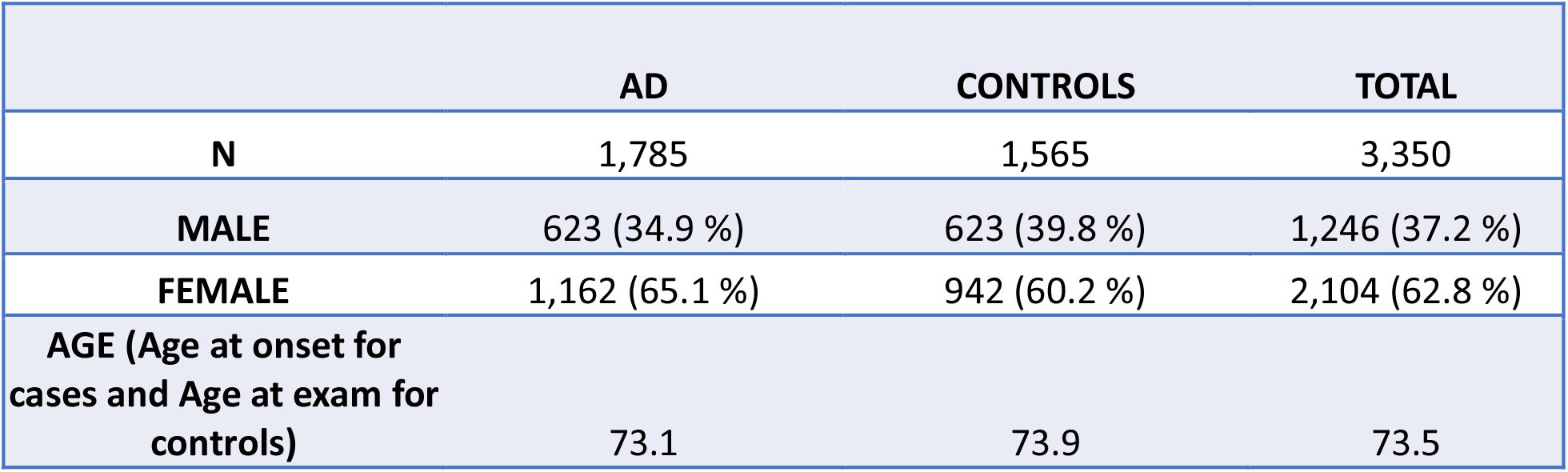
Demographic characteristics of NIA-LOAD data set used for replication. About 53% of the sample was clinically diagnosed with LOAD. Age at onset for AD cases was similar to age at exam for controls. The group of AD has a higher proportion of women (65 vs 60%).

|  | AD | CONTROLS | TOTAL |
| --- | --- | --- | --- |
| **N** | 1,785 | 1,565 | 3,350 |
| **MALE** | 623 (34.9 %) | 623 (39.8 %) | 1,246 (37.2 %) |
| **FEMALE** | 1,162 (65.1 %) | 942 (60.2 %) | 2,104 (62.8 %) |
| **AGE (Age at onset for cases and Age at exam for controls)** | 73.1 | 73.9 | 73.5 |

### Genotyping

DNA extracted from peripheral blood samples was used for genotyping on the Illumina Infinium Global Screening (GSA) or Expanded Multi-Ethnic Genotyping (MEGAex) Array. After pre-imputation QC, which included removing markers missing more than 2% of the data, monomorphic markers, markers that had MAF < 0.05 and markers that failed Hardy Weinberg exact test at P value < 10^−6^, the MEGAex final set before imputation was 552,801 markers and the GSA was 279,931. The two datasets were imputed separately using the Haplotype Reference Consortium (HRC) reference panel. Post-Imputation QC parameters were MAF >= 0.01, r^2^ >=0.4; MAF < 0.01, r^2^ >=0.8. Subsequently, the overlapping markers that passed QC from both sets were merged for a total of over 7 million common and rare imputed variants. Hardy-Weinberg Equilibrium (HWE) was tested for all SNPs and variants significantly out of HWE (P value < 10^−6^) were removed before the analysis. Supplementary Table 1 summarizes the imputed SNPs retained for analysis by chromosome.

The NIA-LOAD dataset was genotyped using the Illumina 610 array containing 592,532 SNPs. For genotyping details see Wijsman et al. (2011). After genotype chip QC, the dataset was imputed to the 1,000 Genomes Project using IMPUTE2. The total number of imputed SNPs before QC was 39,127,740. For imputation details see Kunkle et al. (2019). We concentrated the replication analysis on chromosomes 6, 9 and 17. Supplementary Table 2 shows the number of imputed SNPs analyzed on each chromosome.

### Statistical Analysis

The Cox proportional hazards model is widely used in the analysis of time-to-event phenotypes (such as AAO) and mixed-effects models are commonly used in family-based genetic studies to adjust for relatedness of individuals. We used Coxmeg (He & Kulminski. 2020) an R package for conducting GWAS of age-at-onset traits in related individuals using Cox mixed-effects models (CMEMs). The CU survival time outcome was defined as AAO of CI (cases) or age at last normal cognitive examination (CU (censored) controls) in the Amish and NIA-LOAD (replication) cohorts. Familial relationships were controlled by including a random effect specified by the genetic relationship matrix (GRM) estimated from the genotype data in both Amish and NIH-LOAD datasets. In both datasets sex was included as a covariate. Individual SNPs, coded additively as imputed dosages of the less common allele, were added to this base model to test for association with CU survival time. The strength of this association is estimated by the hazard ratio (HR). The qqman package in R was used to summarize the significance of the association of each SNP with CU survival time in a Manhattan plot of p-values from the chi-square test of association. In the Amish dataset, genome-wide significant results were determined using the traditional GWAS threshold P < 5 × 10^−8^, and suggestive results were highlighted using the threshold P < 1 × 10^−5^. LocusZoom (Pruim et al., 2010) was used to generate regional association plots and Linkage Disequilibrium analysis (LD) of all common variants located in a 1Mb interval centered on SNPs with genome-wide significant associations. Three genome-wide significant results were identified, and markers in a 1-Megabase (Mb) region centered on each peak SNP were selected for analysis in the NIA-LOAD replication dataset, using identical statistical models to the Amish discovery dataset. To assess potential interaction of genome-wide significant effects with *APOE* genotype, stratified analysis was conducted in Amish and NIA-LOAD samples subdivided into APOE-4 carriers and non-carriers.

## RESULTS

### Demographic statistics and genotype quality control for Amish dataset

The demographic characteristics of the Amish dataset are shown in Table 1. A total of 362 subjects were cognitively impaired (CI). Of these, 58% were women. The remaining 1,160 were cognitively unimpaired (CU). Of these, 58.4% were women. Age at last examination was 81.6 for the CI group and 70.6 for the CU group. The mean age at first exam below 3MS cutoff point for the CI group was 81.2 years.

The total number of imputed variants after QC, including rare variants, was 7,819,581 (Supplementary Table 1). Tests of HWE identified 3,630 variants significantly deviating from HWE (p-value < 10^−6^). After removing these variants, 7,815,951 were retained for GWAS analysis.

### GWAS of CU survival time

The GWAS of CU survival time used a Cox mixed-effects model with each imputed variant and sex as a fixed-effects covariate, with the kinship matrix included as a random effect. The results for all autosomes are summarized in the Manhattan plot in Figure 1. Three common variants (minor allele frequency (MAF) > 0.01) were significantly associated (P < 5 × 10^−8^) with CI-free survival time (Table 2) on chromosomes 6 (rs145348074), 9 (rs534551495), and 17 (rs146729640). Table 3 lists for each associated region the p value, hazard ratio, minimum allele frequency (MAF), and overlapping or closest gene and genes located within +/- 500 thousand base pairs. To determine if there were broad regions of association surrounding these peak markers, we examined regional association plots (Supplemental Figures 1-3), identified variants with suggestive evidence of association (p < 10^−5^, Supplemental Table 3), and examined linkage disequilibrium patterns in the region to assess whether there was evidence for multiple association signals.

**Figure 1.**
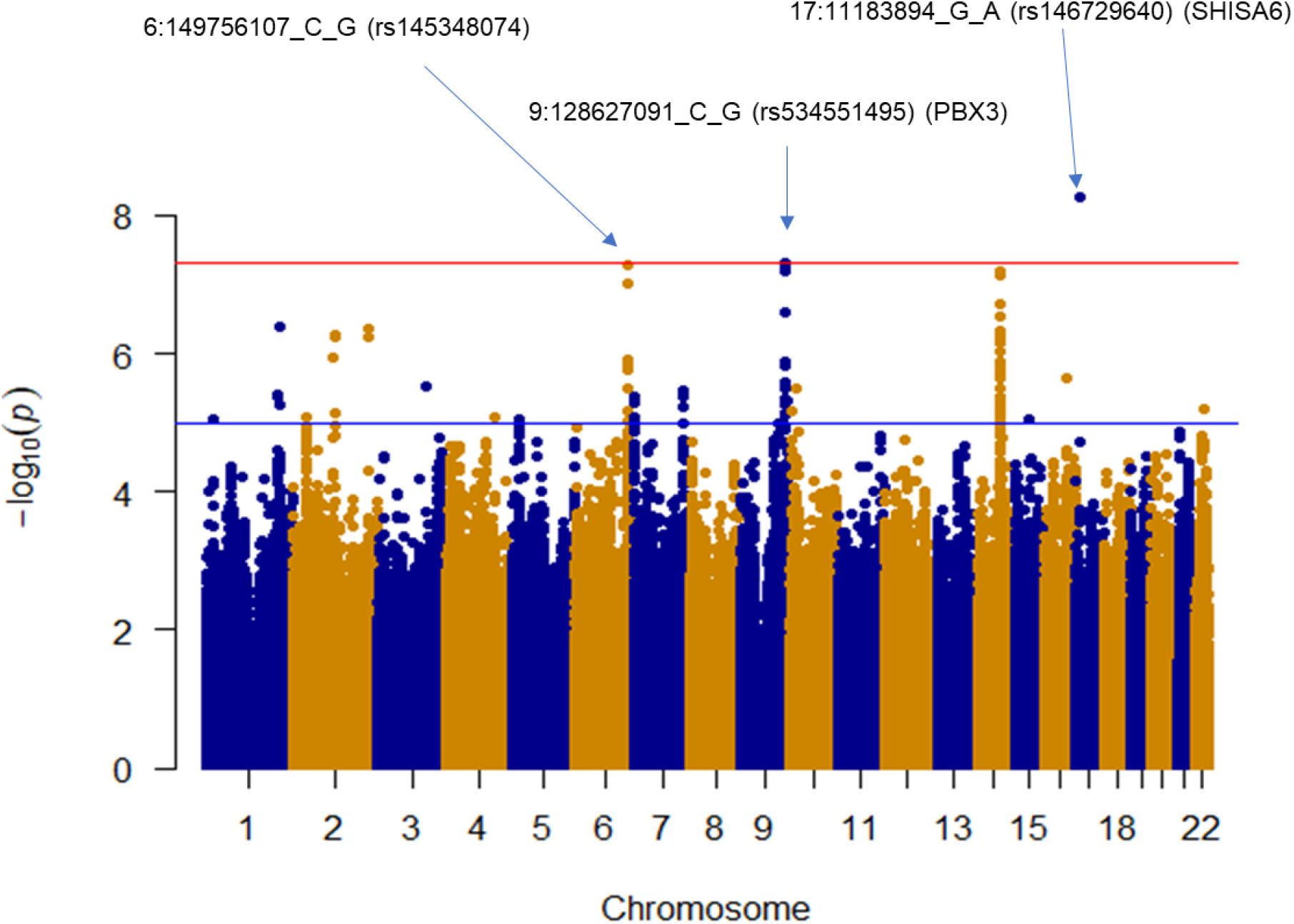
Manhattan plot showing results of GWAS of CU survival time using Cox mixed-effects model: Red line indicates the threshold for genome wide significance (P < 5 × 10^−8^) while the blue line represents the suggestive threshold (P < 1 × 10^−5^). Three variants reached genome wide significance: 6:149756107_C_G (rs145348074), 9:128627091_C_G (rs534551495), and 17:11183894_G_A (rs146729640). Mapping is based on GRCh37/hg19 assembly.

**Table 3.** Three common variants were significantly associated (p < 5 x 10 ^-8^) with CU survival time in the Amish dataset. The strongest result was on chromosome 17, followed by chromosomes 6 and 9. Variants are annotated as overlapping a gene if located within the gene boundaries. Other genes within a 1Mb interval centered on the associated variant are also listed.

| Location and base pair change (build hg19) | SNP (rs) | MAF | Variant type | Hazard Ratio | P-value | Overlapping Genes | Genes within +/- 500Kb |
| --- | --- | --- | --- | --- | --- | --- | --- |
| 6:149756107_C_G | rs145348074 | 0.0237382 | Intergenic | 3.35 | 5.26E-08 | Intergenic variant.<br>Closest gene: *ZC3H12D* | *UST, UST-AS1, LOC105378047, TAB2, TAB2-AS1, SUMO4, ZC3H12D, PPIL4, GINM1, KATNA1, LATS1, LOC645967, NUP43, PCMT1, LRP11, RAET1E-AS1, RAET1E, RAET1G, LOC105378052* |
| 9:128627091_C_G | rs534551495 | 0.0379333 | Intron | 2.82 | 4.83E-08 | *PBX3* | *GAPVD1, MAPKAP1, LOC51145, PBX3, LOC10192911, MVB12B* |
| 17:11183894_G_A | rs146729640 | 0.0119911 | Intron | 6.38 | 5.59E-09 | *SHISA6* | *TMEM220-AS1, DNAH9, TMEM238L, PIRT, SHISA6,* |

### Regional plot and local linkage disequilibrium (LD) analysis for each genome wide variant

As summarized in Table 4, regional association analysis of 1-Megabase (Mb) regions on chromosomes 6 and 9 identified three additional variants in moderate to strong LD (r^2^>0.4) with the peak markers and suggestive evidence of association with AAO (P < 1 ⨯ 10^−5^). No additional markers were identified in the chromosome 17 region. Regional association plots are provided in Supplemental Figures 1, 2 and 3.

**Table 4.**
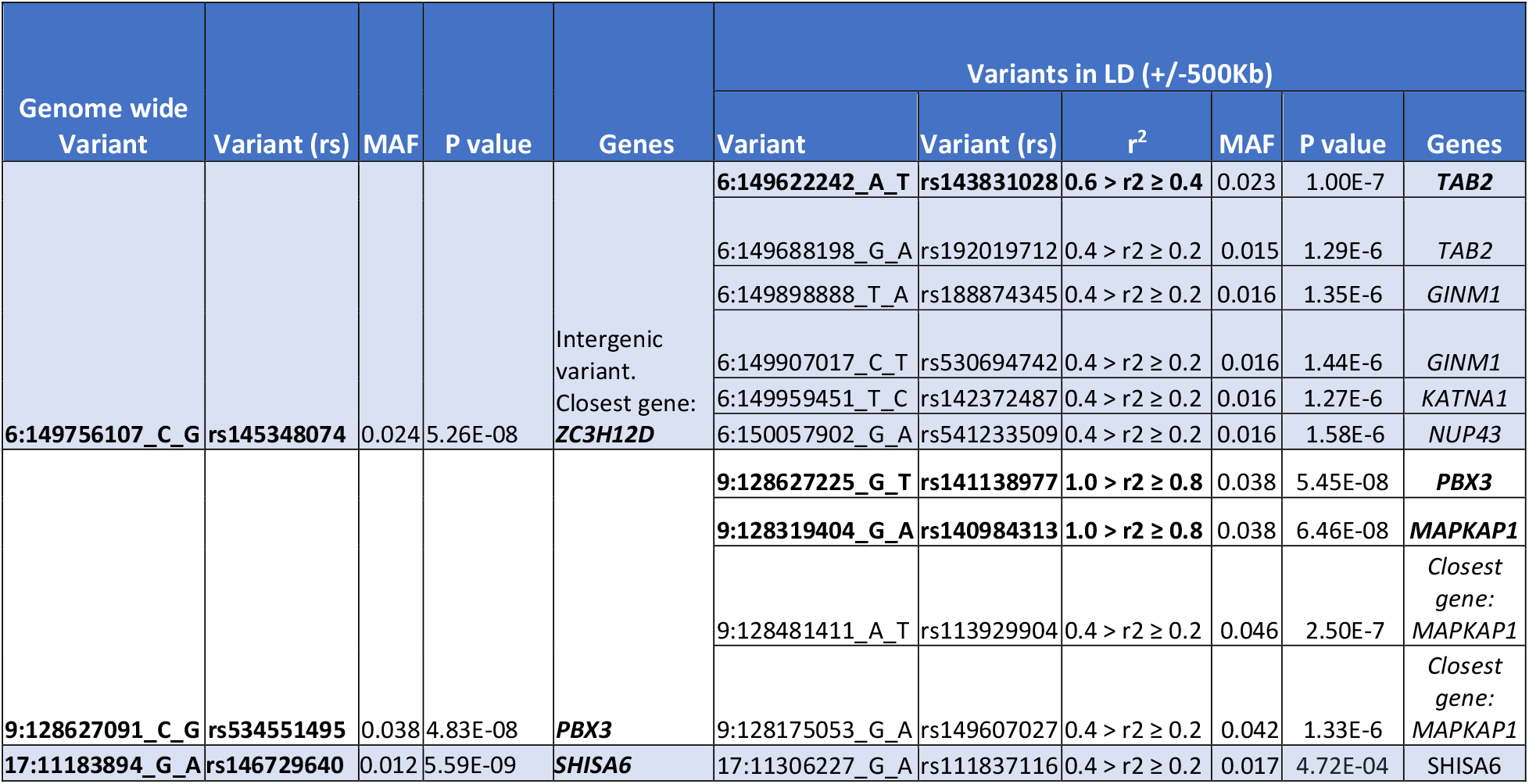
Results of local linkage disequilibrium (LD) analysis of genome wide significant variants. Variants rs141138977 and rs140984313 were found in LD (1.0 > r^2^ ≥ 0.8) with rs534551495, the genome wide signal found on chromosome 9. Variant rs143831028 was in LD (0.6 > r^2^ ≥ 0.4) with rs145348074, the genome wide signal found on chromosome 6. None of the variants was found in LD (r^2^ ≥ 0.4) with rs146729640, the genome wide signal found on chromosome 17.

### Replication analysis of the three genome-wide significant results in the independent NIA-LOAD dataset

The demographic characteristics of the NIA-LOAD replication dataset are shown in Table 2. There are 1,785 people with AD (65% women) and 1,565 CU controls (60% women). The age at onset was 73.1 for cases and the age at exam for the control group was 73.9. The replication analyses were conducted in the 1Mb interval centered on each peak variant from the Amish GWAS. For chromosome 6, variants numbered 14,532; for chromosome 9, variants were 13,602; for chromosome 17 variants were 17,000 (Supplementary Table 2).

Analysis of CU survival time in the NIA-LOAD dataset detected nominal evidence of association in the chromosome 17 interval, at the same peak marker as the Amish GWAS (rs146729640; p=0.02, Table 5). Results for the other two regions were not significant. While the result on chromosome 17 did not exceed the Bonferroni-adjusted significance threshold (p=0.05/17,000 variants tested = 3 × 10^−6^), the replication of the result at the *exact same variant* with nominal significance suggests that the observed association is reproducible.

**Table 5.** Results of the Cox mixed effect model for replication using NIA-LOAD data set show nominal association (p = 0.02) in the chromosome 17 variant (rs146729640).

| Chr | Variant | Variant (rs) | Data set | # of subjects | MAF | Hazard ratio | P-value |
| --- | --- | --- | --- | --- | --- | --- | --- |
| 17 | 17:11183894_G_A | rs146729640 | Amish | 1520 | 0.012 | 6.38 | 5.59E-09 |
|  |  |  | NIA-LOAD | 3350 | 0.012 | 1.49 | 0.0217 |
| 9 | 9:128627091_C_G | rs534551495 | Amish | 1520 | 0.038 | 2.82 | 4.83E-08 |
|  |  |  | NIA-LOAD | 3350 | 0.006 | 0.98 | 0.9356 |
| 6 | 6:149756107_C_G | rs145348074 | Amish | 1520 | 0.024 | 3.35 | 5.26E-08 |
|  |  |  | NIA-LOAD | 3350 | 0.013 | 1.16 | 0.4164 |

### Stratified Analysis of Chromosome 17 regions by APOE-4 carrier status

To explore the possibility that *APOE* genotype might influences these results, the discovery and replication datasets were stratified by APOE-4 carrier status and the Cox mixed-effects models were run in each subset. In both the Amish (Table 6) and NIA-LOAD (Table 7) datasets, the effect (hazard ratio) at rs146729640 was stronger in the subset of individuals that did not carry the APOE-4 allele.

**Table 6.**
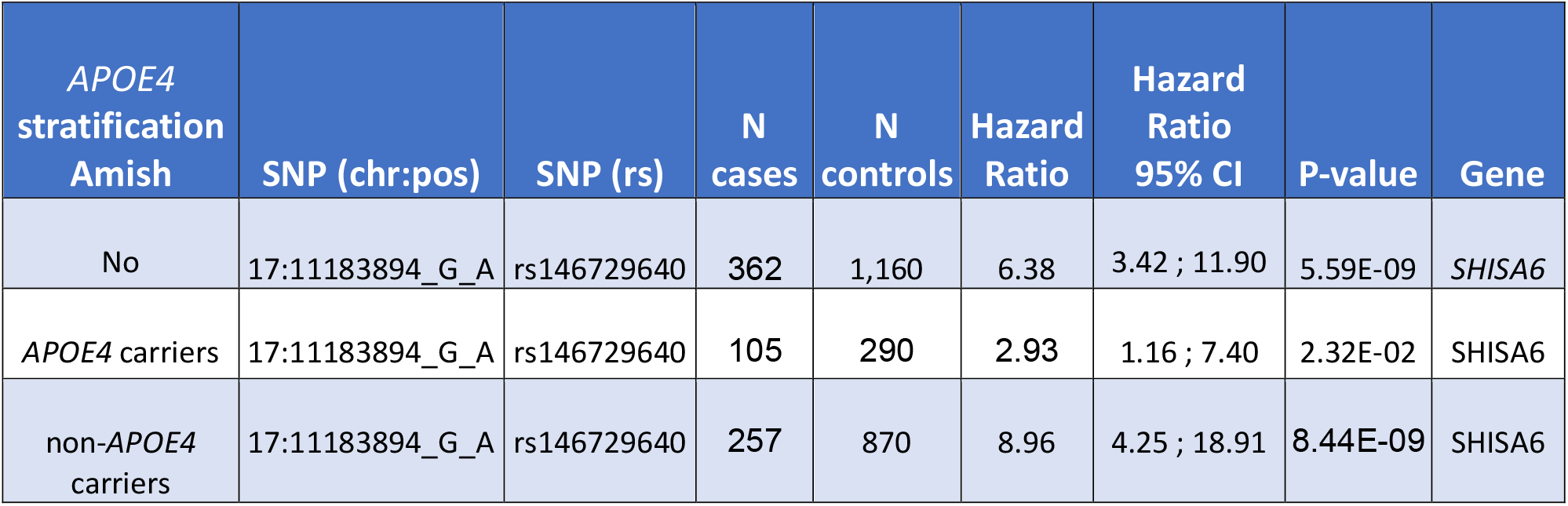
Stratified survival analysis of rs146729640 variant in the Amish dataset. The strength of association was increased in the non- *APOE4* carriers.

**Table 7.** Stratified survival analysis of rs146729640 variant in the NIA-LOAD dataset. Similar to the Amish discovery set, the strength of association was increased in the non-*APOE4* carriers.

| *APOE4*<br>stratification<br>NIA-LOAD | SNP (chr:pos) | SNP (rs) | N<br>cases | N<br>controls | Hazard<br>Ratio | Hazard<br>Ratio<br>95% CI | P-value | Gene |
| --- | --- | --- | --- | --- | --- | --- | --- | --- |
| No | 17:11183894_G_A | rs146729640 | 1,785 | 1,565 | 1.49 | 1.06 ; 2.08 | 0.0217 | SHISA6 |
| *APOE4* carriers | 17:11183894_G_A | rs146729640 | 1326 | 572 | 1.35 | 0.95 ; 1.93 | 0.0950 | SHISA6 |
| non-*APOE4*<br>carriers | 17:11183894_G_A | rs146729640 | 459 | 993 | 1.82 | 0.96 ; 3.43 | 0.0658 | SHISA6 |

## DISCUSSION

This study identified a reproducible association of CU survival time with an intronic polymorphism of the *SHISA6* gene on chromosome 17, where the more frequent allele is associated with later AAO of CI. Large-scale genome-wide association studies (GWAS) of disease-free survival offer a powerful method of identifying factors that delay the onset of disease, manifesting in later AAO. Such discoveries might illuminate mechanisms underlying disease progression of age-related diseases such as CI and AD. In the present study we performed a GWAS of 7.8 M imputed variants, examining association with CU survival time in 1,520 Amish individuals longitudinally evaluated for CI. Three genome-wide significant results were examined for replication in the independent NIA-LOAD family sample of 1785 individuals with AD and 1565 CU controls, and the association at rs145348074 (chromosome 17) was replicated at a nominal significance level in the NIA-LOAD dataset. We note that the outcome of interest in both datasets, time free of cognitive impairment, was the same, despite differences in the definition used to identify CI. The association at rs145348074 in both datasets despite these differences is encouraging. This intronic polymorphism is located within the *SHISA6* gene, an excellent candidate gene for AD.

*SHISA6* has previously been implicated in AD pathogenesis in human proteomics and transgenic mouse model studies. The *SHISA6* is among the 60 proteins, out of 4,582 analyzed, found significantly altered by increasing age in the human hippocampus. *SHISA6* was found downregulated in this study, which was carried out using hippocampal samples from 16 Chinese brain tissue donors free of neurological diseases across four age groups using the youngest one (22-49) as reference: 22–49, 50–69, 70–89, and >90 (Xu et al. 2016). Additionally, several experiments *in vivo* using *APOE* transgenic mice have revealed that the Shisa6 gene is differentially expressed (downregulated log2 fold change = - 0.53) in the entorhinal cortex (EC) of aged *APOE* mice (*APOE3/4 vs. APOE3/3*) (Area-Gomez et al. 2020). Based on these findings, we performed a stratified analysis and found that the strength of association at rs145348074 (*SHISA6* gene) was increased in the non-*APOE4* carriers in both the Amish and the NIA-LOAD replication data sets. Notably, the EC is one of the first regions to be affected by AD pathology in humans. Shisa6 is also involved in maintenance of high-frequency synaptic transmission between neurons in the mouse hippocampus by regulating AMPA-type glutamate receptors (AMPAR) (Klaassen el al. 2016). One limitation in our study is that for people in the Amish cohort who were CI at baseline, the AAO of CI we used (age at exam) was probably later than the true AAO, as symptoms may have started earlier and been unrecognized or unreported prior to examination. This could have biased our results towards the null; therefore, the true association may be stronger than the current estimate.

GWAS studies are susceptible to false positives results due to the large number of statistical tests. Therefore, an internally replicated design that focuses on signals that are independently detected in a second dataset provides stronger evidence of association. While our GWAS was conducted in a family-based study of 1,520 Amish, we tested our three genome-wide significant findings in a second family dataset (NIA-LOAD) with twice the sample size. The replicated result in the *SHISA6* gene is bolstered by its biological plausibility; this gene has been implicated in transgenic mouse models of AD and human proteomic studies of the aging hippocampus. The effect in the NIA-LOAD is somewhat smaller, but still statistically significant. These results illustrate the utility of using isolated populations to facilitate replicable gene discoveries in complex traits. These specific results suggest that *SHISA6* might modulate the rate of cognitive decline, and studies to elucidate the specific effect and mechanism underlying this reproducible association are warranted.

## Supporting information

STROBE Checklist

## Data Availability

Genotype and case/control phenotype data for the NIA/LOAD dataset is deposited in the NIAGADS portal. The genotype and phenotype data from the Amish discovery sample will be deposited in this portal upon publication.

http://www.niagads.org

## ACKNOWLEDGEMENTS

We thank the Amish families for allowing us into their communities and for participating in our study. We also acknowledge the contributions of the Anabaptist Genealogy Database and Swiss Anabaptist Genealogy Association. This study is supported by National Institutes of Health / National Institute of Aging, grant 1RF1AG058066 (to Jonathan L. Haines, Margaret Pericak-Vance, and William K. Scott) and grant U24 AG056270 (to Tatiana M. Foroud, Alison M. Goate and Richard P. Mayeux). Finally, we acknowledge the resources provided by the Department of Population and Quantitative Health Sciences, School of Medicine at Case Western Reserve University and the John P. Hussman Institute for Human Genomics at University of Miami, Miller School of Medicine.

