## Supplemental Figures and Tables for "Genetic variants in the *SHISA6* gene are associated with delayed cognitive impairment in two family datasets"

(1)John P. Hussman Institute for Human Genomics, University of Miami Miller School of Medicine, Miami, FL, USA, (2)Case Western Reserve University School of Medicine, Cleveland, OH, USA, (3)University Hospitals Cleveland Medical Center, Cleveland, OH, USA, (4)The Dr. John T. Macdonald Foundation Department of Human Genetics, University of Miami Miller School of Medicine, Miami, FL, USA, (5) Cleveland Institute for Computational Biology, Case Western Reserve University, Cleveland, OH, USA, (6) Indiana Alzheimer's Disease Center, Indiana University School of Medicine, Indianapolis, IN, USA, (7) Taub Institute on Alzheimer's Disease and the Aging Brain, Department of Neurology, Columbia University, New York, NY, USA, (8) Gertrude H. Sergievsky Center, Columbia University, New York, NY, USA, (9) Department of Neurology, Columbia University, New York, NY, USA, (10) University of Toronto, Toronto, ON, Canada.

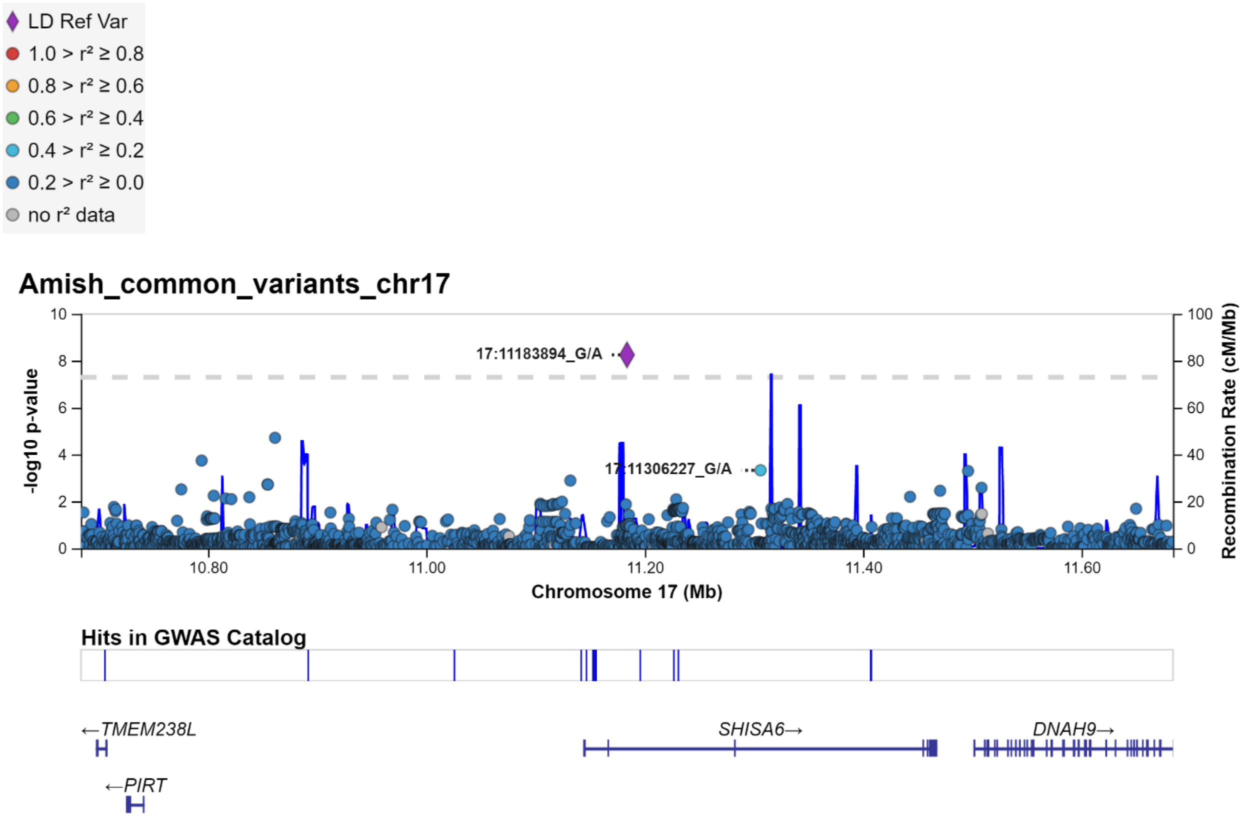

**Supplementary Figure 1. Regional plot of the newly identified genome wide locus 17:11183894_G_A showing the common variants within +/- 500Kb :** The y axis shows the $-\log_{10}$ P values of SNP associations, and the x axis shows their chromosomal positions. Linkage disequilibrium analysis did not reveal any variant in linkage disequilibrium ($r^2 \geq 0.4$) . Color codes for referenced variant and $r^2$ are shown on the bottom of this graph and are equally valid for supplementary figures 2 and 3.

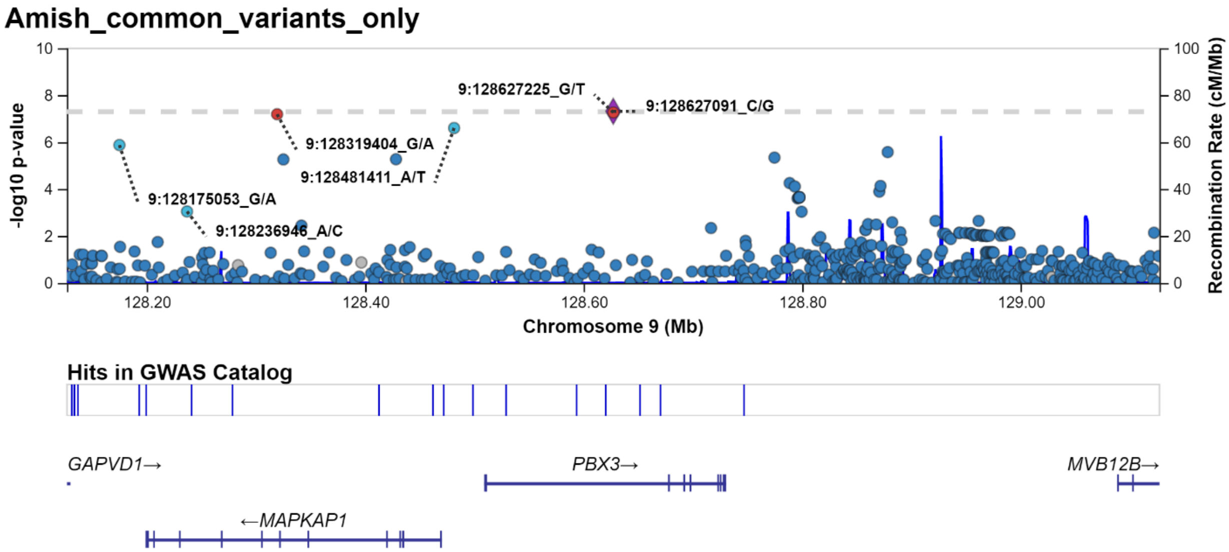

**Supplementary Figure 2. Regional plot of the newly identified genome wide locus 9:128627091_C_G showing the common variants within +/- 500Kb :** The y axis shows the $-\log_{10}$ P values of SNP associations, and the x axis shows their chromosomal positions. Linkage disequilibrium analysis revealed two variants in linkage disequilibrium with the genome wide signal plotted as a purple diamond: 9:128627225_G_T and 9:128319404_G_A. Both have similar r values = 1.0 > r2 ≥ 0.8 and are plotted in red.

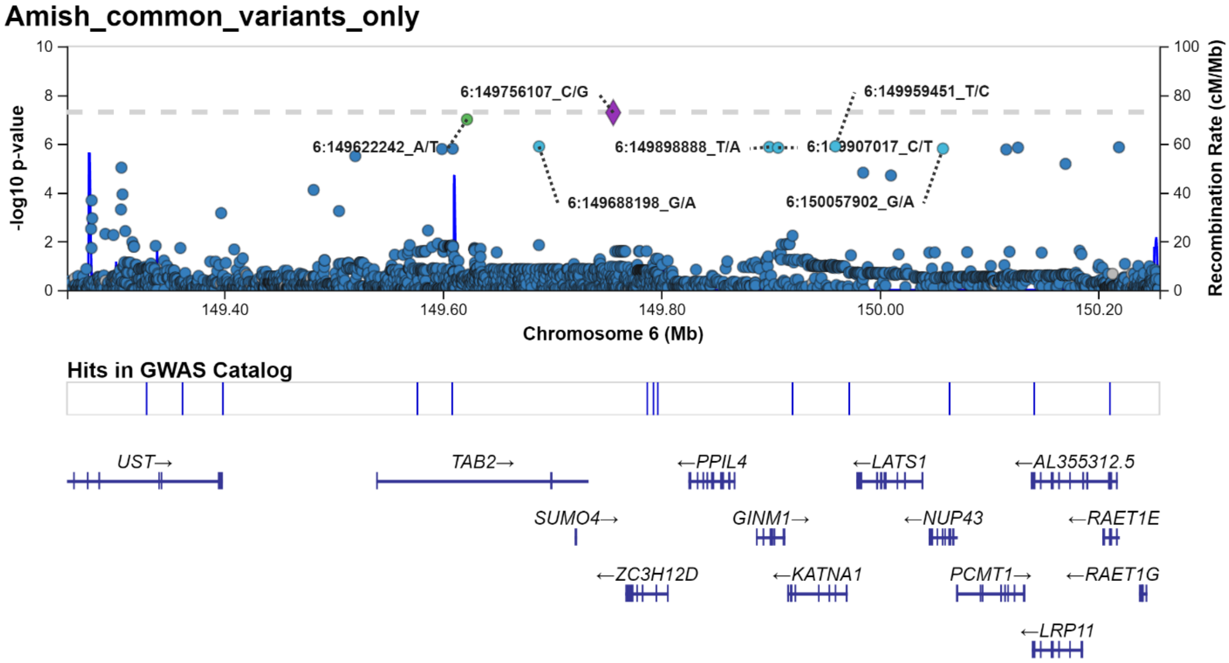

**Supplementary Figure 3. Regional plot showing the newly identified genome wide locus 6:149756107_C_G showing the common variants within +/- 500Kb :** The y axis shows the $-\log_{10}$ P values of SNP associations, and the x axis shows their chromosomal positions. Linkage disequilibrium analysis revealed one variant in linkage disequilibrium with the genome wide signal plotted as a purple diamond: 6:149622242_A_T with r value = 0.6 > r2 ≥ 0.4 and plotted in green.

**Supplementary Table 1. Imputed variants used in the Cox mixed effect model in the Amish cohort.**

| Chromosome | Number of variants | Variants removed HWE test p<10E-06 | Number of variants for survival analysis |
|---|---|---|---|
| chr1 | 594,785 | 193 | 594,592 |
| chr2 | 662,547 | 243 | 662,304 |
| chr3 | 558,312 | 289 | 558,023 |
| chr4 | 576,920 | 188 | 576,732 |
| chr5 | 511,803 | 173 | 511,630 |
| chr6 | 527,109 | 238 | 526,871 |
| chr7 | 458,448 | 165 | 458,283 |
| chr8 | 435,690 | 481 | 435,209 |
| chr9 | 337,624 | 149 | 337,475 |
| chr10 | 399,891 | 151 | 399,740 |
| chr11 | 389,580 | 280 | 389,300 |
| chr12 | 377,283 | 105 | 377,178 |
| chr13 | 292,304 | 198 | 292,106 |
| chr14 | 257,425 | 37 | 257,388 |
| chr15 | 218,285 | 77 | 218,208 |
| chr16 | 241,366 | 74 | 241,292 |
| chr17 | 200,339 | 207 | 200,132 |
| chr18 | 224,492 | 147 | 224,345 |
| chr19 | 169,621 | 122 | 169,499 |
| chr20 | 175,073 | 60 | 175,013 |
| chr21 | 106,110 | 36 | 106,074 |
| chr22 | 104,574 | 17 | 104,557 |
| **Total** | 7,819,581 | 3,630 | **7,815,951** |

**Supplementary Table 2. Imputed variants included in replication analysis of CU survival time in the NIA-LOAD dataset.**

| Chr. | Number of HRC imputed variants | Variants removed HWE test p<10E-06 | Number of variants after HWE removal | N of variants in the segment +/- 500 Kb of the variant of interest used for replication of survival analysis |
|---|---|---|---|---|
| chr6 | 2,460,116 | 664 | 2,459,452 | 14,532 |
| chr9 | 1,686,472 | 290 | 1,686,182 | 13,602 |
| chr17 | 1,090,074 | 125 | 1,089,949 | 17,000 |

**Supplementary Table 3. Variants (n=117) with suggestive evidence of association (0.00000005<p<0.00001) with CU survival time in the Amish discovery dataset.**

| Chromosome | Position (hg19) | Chromosome:SNP Position_REF_ALT | P-value | Minor allele frequency |
|---|---|---|---|---|
| 1 | 20140581 | 1:20140581_C_T | 8.72E-06 | 0.031651 |
| 1 | 208458894 | 1:208458894_C_T | 4.32E-06 | 0.014727 |
| 1 | 208477213 | 1:208477213_T_C | 4.03E-06 | 0.014705 |
| 1 | 209602593 | 1:209602593_G_A | 5.5E-06 | 0.015288 |
| 1 | 209694870 | 1:209694870_G_A | 4.12E-07 | 0.020984 |
| 2 | 40770839 | 2:40770839_C_T | 8.39E-06 | 0.026365 |
| 2 | 40771905 | 2:40771905_A_C | 8.36E-06 | 0.026369 |
| 2 | 40772148 | 2:40772148_C_G | 8.36E-06 | 0.026369 |
| 2 | 40772460 | 2:40772460_C_A | 8.43E-06 | 0.026364 |
| 2 | 40772544 | 2:40772544_T_C | 8.36E-06 | 0.026371 |
| 2 | 40773299 | 2:40773299_C_G | 8.37E-06 | 0.026357 |
| 2 | 40773436 | 2:40773436_A_G | 8.37E-06 | 0.02624 |
| 2 | 118618295 | 2:118618295_G_C | 1.17E-06 | 0.01494 |
| 2 | 118700178 | 2:118700178_A_G | 5.55E-07 | 0.015795 |
| 2 | 118741278 | 2:118741278_G_A | 5.82E-07 | 0.015141 |
| 2 | 119116229 | 2:119116229_T_C | 7.5E-06 | 0.024405 |
| 2 | 217071588 | 2:217071588_A_G | 5.93E-07 | 0.0157 |
| 2 | 217130712 | 2:217130712_T_C | 4.4E-07 | 0.014714 |
| 2 | 217137417 | 2:217137417_T_A | 4.39E-07 | 0.014713 |
| 3 | 138927043 | 3:138927043_G_A | 2.93E-06 | 0.013136 |
| 4 | 140170092 | 4:140170092_C_A | 8.65E-06 | 0.288449 |
| 5 | 19735430 | 5:19735430_T_C | 8.8E-06 | 0.218936 |
| 6 | 149305906 | 6:149305906_G_A | 9.5E-06 | 0.201378 |
| 6 | 149519973 | 6:149519973_T_C | 3.2E-06 | 0.02827 |
| 6 | 149599306 | 6:149599306_A_C | 1.64E-06 | 0.071362 |
| 6 | 149609189 | 6:149609189_G_A | 1.59E-06 | 0.071147 |
| 6 | 149622242 | 6:149622242_A_T | 1E-07 | 0.023211 |
| 6 | 149688198 | 6:149688198_G_A | 1.29E-06 | 0.01481 |
| 6 | 149898888 | 6:149898888_T_A | 1.35E-06 | 0.016102 |
| 6 | 149907017 | 6:149907017_C_T | 1.44E-06 | 0.016293 |
| 6 | 149959451 | 6:149959451_T_C | 1.27E-06 | 0.0157 |
| 6 | 150057902 | 6:150057902_G_A | 1.58E-06 | 0.016012 |
| 6 | 150115865 | 6:150115865_C_T | 1.71E-06 | 0.016344 |
| 6 | 150126590 | 6:150126590_C_T | 1.43E-06 | 0.015804 |
| 6 | 150170088 | 6:150170088_G_A | 6.66E-06 | 0.012037 |
| 6 | 150219019 | 6:150219019_C_T | 1.39E-06 | 0.015236 |
| 6 | 151300032 | 6:151300032_C_A | 3.16E-06 | 0.020878 |
| 7 | 463355 | 7:463355_G_A | 9.36E-06 | 0.486754 |

| | | | | |
|---|---|---|---|---|
| 7 | 475642 | 7:475642_T_C | 4.34E-06 | 0.492762 |
| 7 | 476355 | 7:476355_G_C | 5.34E-06 | 0.475134 |
| 7 | 477735 | 7:477735_T_C | 4.93E-06 | 0.476862 |
| 7 | 477893 | 7:477893_T_C | 8.51E-06 | 0.496056 |
| 7 | 138843420 | 7:138843420_G_T | 3.54E-06 | 0.014549 |
| 7 | 138861291 | 7:138861291_C_T | 5.97E-06 | 0.016992 |
| 7 | 138872420 | 7:138872420_A_G | 5.86E-06 | 0.015474 |
| 7 | 138931615 | 7:138931615_C_T | 4.19E-06 | 0.014073 |
| 9 | 126058380 | 9:126058380_G_A | 7.99E-06 | 0.019233 |
| 9 | 126078275 | 9:126078275_C_A | 7.5E-06 | 0.02163 |
| 9 | 127753308 | 9:127753308_T_C | 3.14E-06 | 0.020707 |
| 9 | 127844665 | 9:127844665_G_A | 1.5E-06 | 0.023712 |
| 9 | 128175053 | 9:128175053_G_A | 1.33E-06 | 0.041516 |
| 9 | 128319404 | 9:128319404_G_A | 6.46E-08 | 0.03754 |
| 9 | 128325149 | 9:128325149_T_C | 5.48E-06 | 0.020272 |
| 9 | 128428222 | 9:128428222_G_C | 5.36E-06 | 0.02032 |
| 9 | 128481411 | 9:128481411_A_T | 2.5E-07 | 0.046468 |
| 9 | 128627225 | 9:128627225_G_T | 5.45E-08 | 0.037619 |
| 9 | 128774677 | 9:128774677_A_G | 4.56E-06 | 0.023603 |
| 9 | 128878311 | 9:128878311_C_T | 2.66E-06 | 0.026973 |
| 9 | 131208443 | 9:131208443_C_T | 4.94E-06 | 0.026841 |
| 10 | 4615800 | 10:4615800_A_G | 6.9E-06 | 0.046516 |
| 10 | 18789075 | 10:18789075_T_C | 3.15E-06 | 0.206483 |
| 14 | 80822381 | 14:80822381_A_G | 9.15E-06 | 0.023931 |
| 14 | 81054839 | 14:81054839_G_A | 8.31E-06 | 0.036802 |
| 14 | 81292975 | 14:81292975_T_C | 9.65E-06 | 0.037061 |
| 14 | 81973297 | 14:81973297_T_C | 2.94E-07 | 0.032668 |
| 14 | 82321005 | 14:82321005_T_C | 1.59E-06 | 0.0254 |
| 14 | 82323777 | 14:82323777_A_G | 9.78E-06 | 0.036224 |
| 14 | 82348283 | 14:82348283_G_A | 7.67E-06 | 0.035386 |
| 14 | 82399291 | 14:82399291_G_A | 6.46E-06 | 0.037457 |
| 14 | 82414238 | 14:82414238_G_C | 5.42E-07 | 0.026538 |
| 14 | 82464419 | 14:82464419_A_G | 3.11E-06 | 0.023973 |
| 14 | 82487131 | 14:82487131_A_G | 1.81E-06 | 0.024065 |
| 14 | 82516708 | 14:82516708_T_G | 1.81E-06 | 0.024068 |
| 14 | 82745357 | 14:82745357_G_C | 1.34E-06 | 0.026061 |
| 14 | 82808245 | 14:82808245_G_A | 1.48E-06 | 0.026906 |
| 14 | 82846606 | 14:82846606_G_A | 8.8E-06 | 0.037754 |
| 14 | 82868633 | 14:82868633_G_A | 6.32E-08 | 0.129653 |
| 14 | 82868809 | 14:82868809_T_C | 7.37E-08 | 0.130428 |
| 14 | 82869446 | 14:82869446_G_A | 1.88E-07 | 0.043562 |

| | | | | |
|---|---|---|---|---|
| 14 | 82875943 | 14:82875943_T_G | 7.11E-08 | 0.131749 |
| 14 | 82876272 | 14:82876272_C_T | 7.09E-08 | 0.131744 |
| 14 | 82878686 | 14:82878686_G_A | 6.66E-08 | 0.131874 |
| 14 | 82909289 | 14:82909289_T_C | 1.45E-06 | 0.025715 |
| 14 | 83144455 | 14:83144455_T_C | 1.63E-06 | 0.025325 |
| 14 | 83188416 | 14:83188416_T_G | 9.47E-07 | 0.038106 |
| 14 | 83190167 | 14:83190167_A_G | 7.11E-07 | 0.026759 |
| 14 | 83190170 | 14:83190170_A_G | 7.11E-07 | 0.026759 |
| 14 | 83190716 | 14:83190716_C_G | 7.09E-07 | 0.026756 |
| 14 | 83191253 | 14:83191253_G_A | 6.55E-07 | 0.02722 |
| 14 | 83191486 | 14:83191486_G_A | 6.35E-07 | 0.02667 |
| 14 | 83195059 | 14:83195059_A_G | 6.52E-07 | 0.026655 |
| 14 | 83197317 | 14:83197317_C_A | 6.23E-07 | 0.026648 |
| 14 | 83198854 | 14:83198854_G_A | 5.38E-07 | 0.026952 |
| 14 | 83199690 | 14:83199690_A_C | 2.24E-06 | 0.025277 |
| 14 | 83200422 | 14:83200422_T_C | 5.31E-07 | 0.026922 |
| 14 | 83200933 | 14:83200933_G_C | 5.31E-07 | 0.026922 |
| 14 | 83201301 | 14:83201301_A_C | 5.31E-07 | 0.026922 |
| 14 | 83201519 | 14:83201519_C_T | 5.31E-07 | 0.026922 |
| 14 | 83202038 | 14:83202038_C_T | 5.31E-07 | 0.026922 |
| 14 | 83203657 | 14:83203657_T_A | 5.31E-07 | 0.026922 |
| 14 | 83204114 | 14:83204114_T_C | 5.12E-07 | 0.026934 |
| 14 | 83204633 | 14:83204633_C_G | 5.14E-07 | 0.026935 |
| 14 | 83206052 | 14:83206052_C_T | 5.12E-07 | 0.026934 |
| 14 | 83207499 | 14:83207499_C_A | 5.14E-07 | 0.026935 |
| 14 | 83210410 | 14:83210410_G_C | 4.84E-07 | 0.026952 |
| 14 | 83211543 | 14:83211543_T_C | 4.84E-07 | 0.026952 |
| 14 | 83211896 | 14:83211896_G_A | 4.84E-07 | 0.026952 |
| 14 | 83213080 | 14:83213080_C_T | 4.84E-07 | 0.026952 |
| 14 | 83516092 | 14:83516092_C_A | 4.19E-06 | 0.030025 |
| 14 | 83522841 | 14:83522841_A_G | 5.47E-06 | 0.023414 |
| 14 | 83608574 | 14:83608574_T_G | 4.66E-06 | 0.021538 |
| 14 | 83681919 | 14:83681919_G_A | 5.62E-06 | 0.021005 |
| 14 | 83708722 | 14:83708722_A_G | 5.53E-06 | 0.020627 |
| 14 | 84552412 | 14:84552412_T_A | 9.94E-06 | 0.017802 |
| 14 | 84552413 | 14:84552413_C_A | 9.97E-06 | 0.017803 |
| 15 | 58600722 | 15:58600722_C_G | 9.19E-06 | 0.34956 |
| 16 | 64412126 | 16:64412126_T_C | 2.28E-06 | 0.228432 |
